# Risk factors for community transmission of SARS-CoV-2. A cross-sectional study in 116,678 people

**DOI:** 10.1101/2020.12.23.20248514

**Authors:** Eyrun F. Kjetland, Karl Trygve Kalleberg, Camilla Lund Søraas, Bato Hammarström, Tor Åge Myklebust, Synne Jenum, Eyvind Axelsen, Andreas Lind, Roar Bævre-Jensen, Silje Bakken Jørgensen, Frank Olav Pettersen, Lene B. Solberg, Cathrine Lund Hadley, Mette S. Istre, Knut Liestøl, John Arne Dahl, Giske Ursin, Arne Søraas

## Abstract

**Background:** The risk factors for SARS-CoV-2 transmission are not well characterised in Western populations. We sought to identify potential risk factors for transmission and actionable information to prevent for SARS-CoV-2.

**Methods:** Individuals tested for SARS-CoV-2 at four major laboratories were invited. In addition, participants were sampled by convenience after a media campaign. Self-reported test results were compared with laboratory results, demographic data and behavioural facts were collected using a digital platform. In a cross-sectional design positive cases were compared with negative and untested control groups.

**Findings:** Approximately 14 days after a countrywide lockdown in Norway, 116,678 participants were included. Median age was 46 years, 44% had children in preschool or in school; 18% were practicing health professionals. International flights, contact with infected, and gatherings of more than 50 people, were associated with high risk. Health professionals who used public transport were at higher risk of testing positive than those who did not. Having undergone light infections, the last six months was strongly associated with lower odds ratio of SARS-CoV-2 positivity. Contact with children, use of hand sanitiser and use of protective gloves in private were also associated with lower odds ratio of testing positive for SARS-CoV-2.

**Interpretation:** Further research is needed to explore if being a parent or looking after children is associated with lower risk of SARS-CoV-2 positivity in the next phases of the pandemic. Immunological research should be done to determine the effects of prior trivial infections on SARS-CoV-2 infection. We confirm that large gatherings during the pandemic should be avoided and those who are infected, or under suspicion thereof, posed very high risks to others this population.

## Introduction

The Severe Acute Respiratory Syndrome Coronavirus-2 (SARS-CoV-2) causes COVID-19, a potentially fatal disease. At the time of writing, more than almost 75 million people are infected and 1.6 million deaths have been reported in 187 countries.^1^ The World Health Organisation (WHO) declared COVID-19 a pandemic March 11^th^ 2020.^2^

In China, where the virus was first detected, a massive lockdown of society lead to control of the number of COVID-19 cases.^3,4^ In South Korea, closure of schools and other protective measures have interrupted viral transmission.^5^ Strategies such as physical distancing, quarantine of exposed individuals such as travellers, school closures, cessation of large gatherings, and closures of restaurants have been implemented in many countries.^6^

Norway was one of the first Western countries to have a substantial outbreak. The population-wide digital literacy makes the country suitable for a secure, large-scale study in pandemic circumstances. With the aim to guide health policy, we sought to explore the community and health professional risk factors for SARS-CoV-2 acquisition.

## Materials and methods

### Study area and population

The study period was January 1^st^ to April 6^th^ 2020. Study participants were based in Norway, which has a population of 5.4 million people. The Gross Domestic Product (GDP) per capita was USD 92,121 in 2018, whilst the European GDP was USD 43,188.^7^ There are 22.4 practicing nurses and doctors per 1,000, which is the highest amongst the OECD countries (Organisation for Economic Co-operation and Development).^8^ There is high computer literacy, universal free health care, and most health care professionals in the capital city region commute to work by public transport. As shown in Figure 1, March 13^th^ the authorities instituted a lockdown which entailed closure of schools, preschools, restaurants, entertainment, and public gatherings, authorities retracted entry visas and dismissed foreigners at the borders. The population was asked to practice two metre physical distancing, work from home and avoid public transport. Eligible study participants were 18 years or older, had a Norwegian identification number, and electronic access to the national two-factor electronic login system; which is used to access all digital government services.

**Figure 1.**
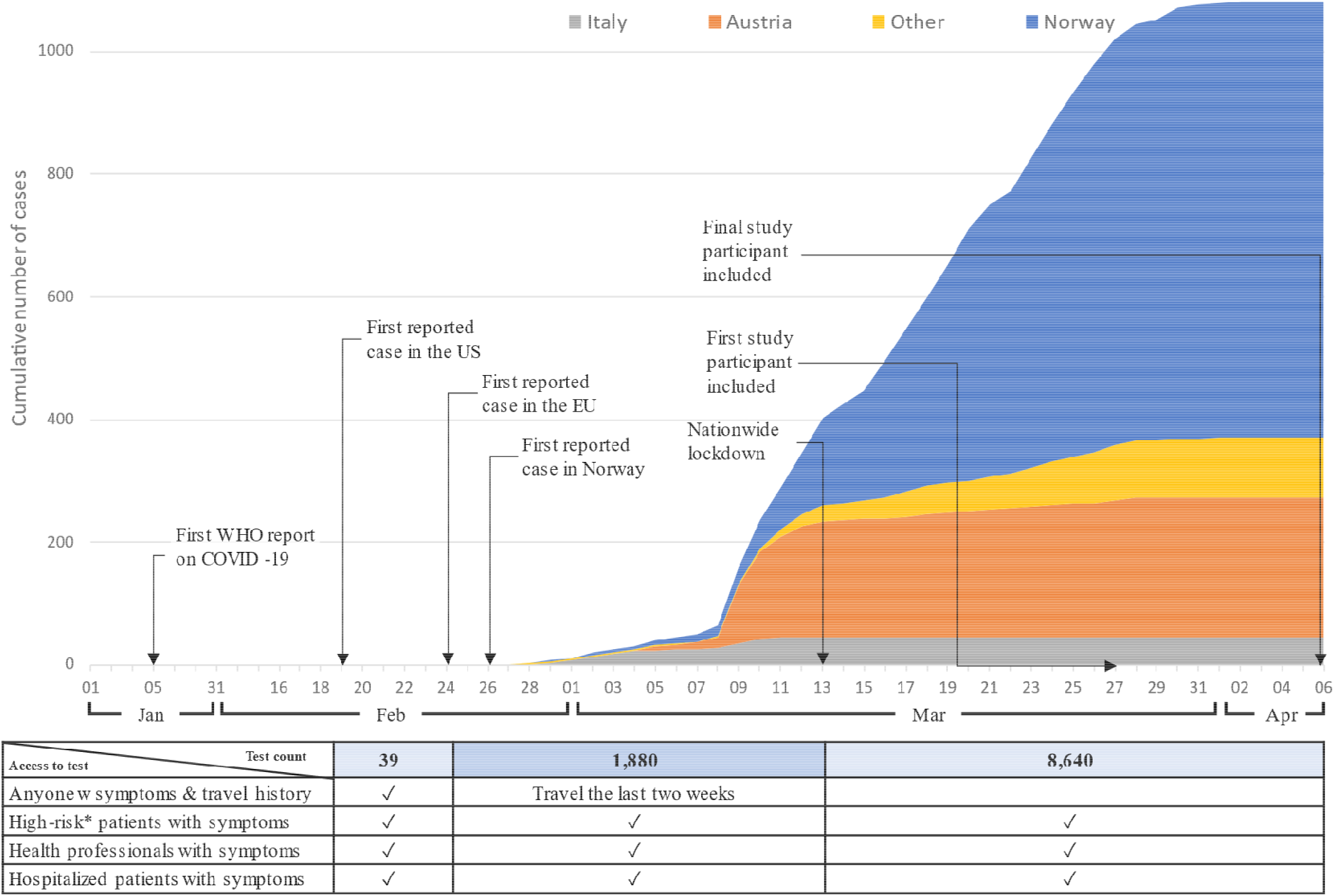
Timeline of the epidemic development in relation to the study population. The countries represent the probable origin of infection. Test criteria changed in the study period.^20,21^ *Above the age of 65 years with cardiovascular or lung disease, cancer, hypertension or diabetes

### Study design and case definition

As shown in Figure 1 the availability of the SARS-CoV-2 PCR test varied in the study period. Figure 2 shows the cross-sectional study design. SARS-CoV-2 positive cases will be compared, firstly with those who had symptoms of COVID-19, but a negative test (denoted negative controls) and, secondly, with those untested who were sampled by convenience (denoted untested volunteer controls). The study was based on two recruitment strategies:

**Figure 2.**
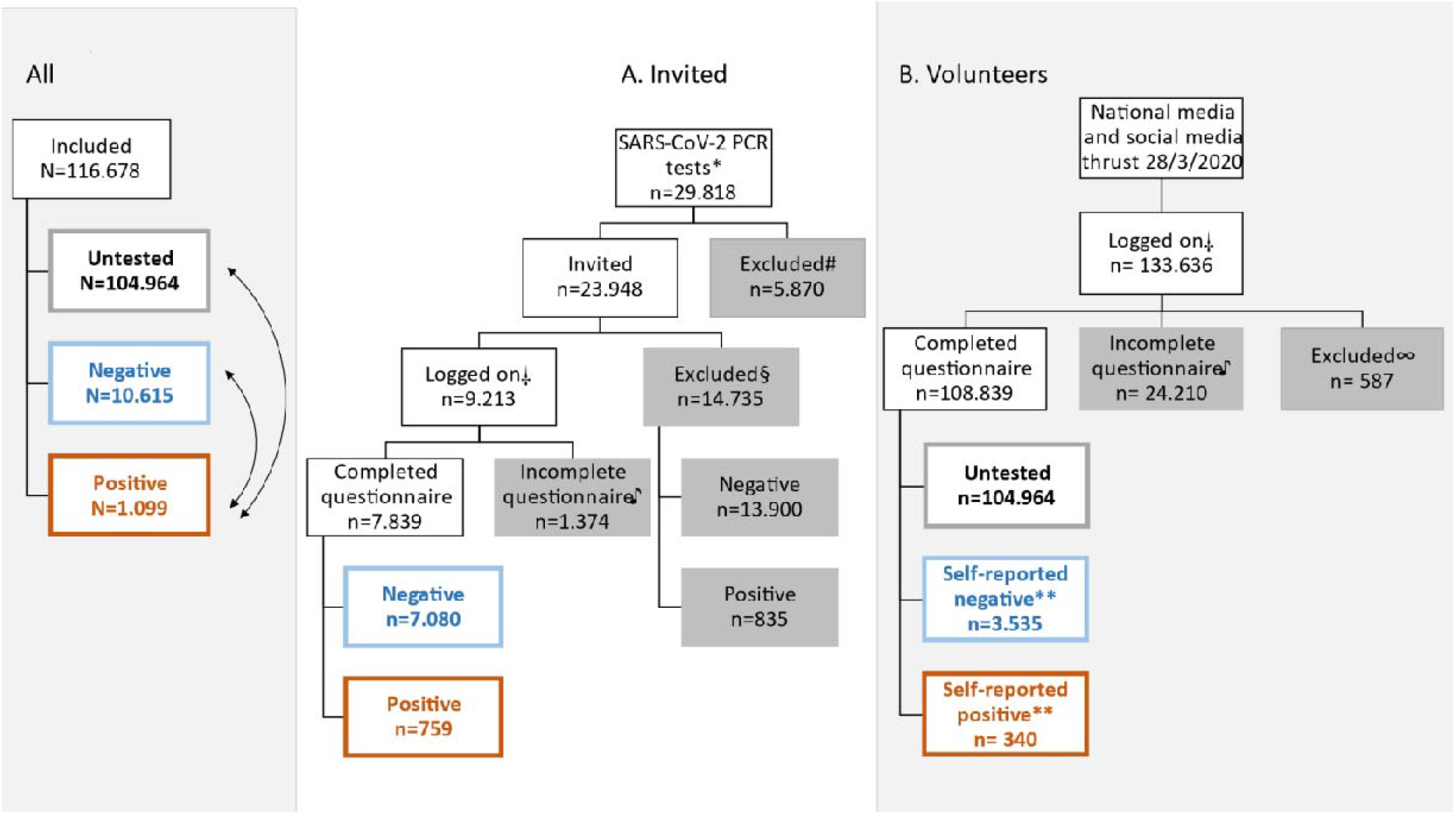
Recruitment of invited (laboratory results) and volunteers (national TV and social media) *Participants tested at Oslo University Hospital, Vestre Viken or Fürst laboratories. #Minors (n=1207), invalid contact information or people in a registry for those who do not want unsolicited contact (n=3276), invalid test results (n=1135), multiple tests from same person (positive test result, n=1681). Some participants had several reasons for exclusion. §Did not respond to invitation before April 6^th^ 2020. □Through online portal. ♪Overload of system. ∞Minors (n=121), refused (n=464), withdrew (n=2). **508 of these could be confirmed, Kappa 0.97.

1. **Invited** (Figure 2A): Between March 27^th^ and April 4^th^ invitations were sent by cellular (cell) phone text message to individuals (n=23,948) who had been tested for SARS-CoV-2 at the Oslo University Hospital, Vestre Viken Hospital, or Fürst laboratories, three of the largest laboratories in and near Oslo. There was high agreement between self-reported and laboratory-reported SARS-CoV-2 PCR results (Kappa 0.99).
2. **Volunteers** (Figure 2B): From March 28^th^, the general population was invited through the Oslo University Hospital Facebook page and the study received nationwide media coverage. Anyone living in Norway was encouraged to participate. As shown in Figure 3 there was high agreement between the geographic study participant distribution and the dissemination of the disease in Norway. Furthermore, there was high agreement between self-reported and laboratory-reported SARS-CoV-2 PCR results (Kappa 0.97) for the 508 tested volunteers for whom we could confirm test results (Supplement 1).

### Data collection

Using a secure digital platform, available through smartphones and computers, participants were asked to complete an online questionnaire on travel history, exposure to known COVID-19 cases, living arrangements, history of disease, self-reported test results and demographic variables. Participants were questioned about their behaviour in the two-week period before the lockdown or, if applicable, before they fell ill/were tested. They were asked about having undergone “light infections” the past six months, in Norway this term implies respiratory tract infections not requiring hospitalisation or antibiotics or cystitis.^9^

**Figure 3.**
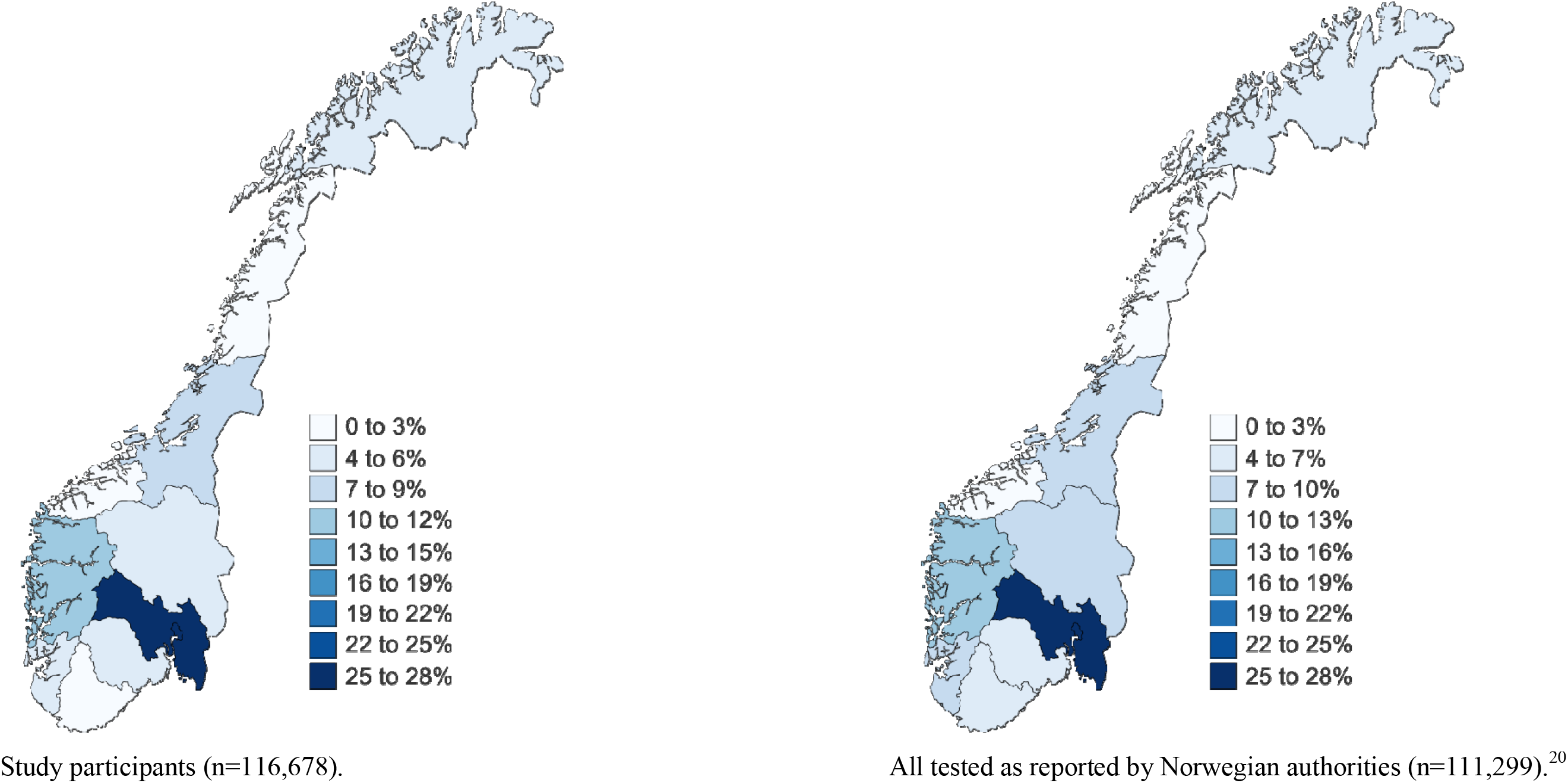
Geographical distribution of study participants and national SARS-CoV-2 cases by April 6, 2020. Information per province by April 6, 2020. The percentages represent the provincial participation and SARS-CoV-2 prevalence.

### SARS-CoV-2 RT-PCR method

SARS-CoV-2 sampling and detection was done according to official national guidelines.^22^ A combined nasopharyngeal and oropharyngeal specimen collected by a health professional and transported to the lab in liquid Amies transport medium where detection of SARS-CoV-2 was done by a reverse transcription polymerase chain reaction (RT-PCR) protocol. All participating laboratories were accredited.

For brevity, only a the Vestre Viken HF method for SARS-CoV-2 reverse transcription polymerase chain reaction (RT-PCR) protocol is included described in detail: Nucleic acid extraction was performed with either MagNA Pure 96 using “MagNA Pure 96 DNA and Viral Small Volume Kit” (Roche) or MagNA Pure LC 2.0 using “MagNA Pure LC Total Nucleic Acid Isolation Kit High Performance” (Roche). RT-qPCR was performed in a duplex assay, including RNase P for internal control on a Lightcycler® 480 II (Roche) targeting the E gene in accordance with Corman et al., using either qScript XLT One-Step RT-qPCR ToughMix (Quantabio) or LightCycler® Multiplex RNA Virus Master (Roche).^23^

### Statistical considerations

Statistical analyses were carried out accordingly. In brief, we compared demographic facts and risk factors between cases and controls. Bivariate and multivariable odds ratios (ORs) and 95% confidence intervals (95% CIs) were estimated as measures of relative risk using logistic regression. Age and sex were considered mandatory confounders and included in all multivariable analyses. A list of potential risk factors was made *a priori* and included in the first round of multivariable analysis. Risk factors with no apparent association with the outcome (ORs very close to 1), were removed from the final model. Statistical significance of the risk factors was assessed using standard likelihood-ratio tests.

In the early phase of the pandemic SARS-CoV-2 PCR tests were widely available for any subject with a suspicion of COVID-19 disease. However, as shown in Figure 1, from March 13^th^, testing was restricted. Only patients potentially requiring hospitalisation, and symptomatic health professionals, were tested. Therefore, separate analyses were done for health professionals. The first cases in Norway had been on ski vacation in Italy or Austria. We consequently performed sensitivity analyses excluding the skiers and those reporting close contact with COVID-19 cases (Supplement 3). No formal mathematical correction was made for multiple comparisons. Data was analysed using SPSS version 26 (IBM Corp, New York, USA) and Stata version 16.0 (Stata Corp LLC, Texas, USA).

### Ethical considerations

The study was approved by the Norwegian ethics committee (REK 124170) and followed the Helsinki Declaration. It was registered in ClinicalTrials.gov (NTC 04320732). All participants were given information about the study, and their right to withdraw from the study at any time. Consent forms were signed electronically, withdrawals/refusals likewise. Data collection and storage was administered through the University of Oslo Services for Sensitive Data.

## Results

In the course of 11 days 116,678 participants were included, 52% were from the greater Oslo region, the epicentre of the epidemic in Norway (Figure 3). Those who had been tested in one of four laboratories of the greater Oslo region represented 6.7% (7839) of the study population. The rest (93.3%) were sampled by convenience as shown in Figure 2B. The time between testing and baseline (inclusion) was on average 13 days (SD=7 days). The cases and two control groups were well matched for age and chronic diseases such as hypertension and diabetes as shown in Table 1. Median age was 46 years (range 19-101), only 3.8% were above the age of 65 years, 71% of the study population (82,948) were female, among negative controls 78% were female (8,497/10,581). There was a median of 3 people (range 1-7) in each household, 44% (51,246) had children, 37,784 in preschool (age 0-5 years) and 38,586 in schools (6 – 19 years), 25,124 had children in both categories. One third lived in apartments, 2-3 % had diabetes, 10-13% reported hypertension, one in five had dogs and almost as many had cats. Health professionals, with and without patient contact, constituted 24% (27,567) of the study participants (Supplement 2).

**Table 1.**
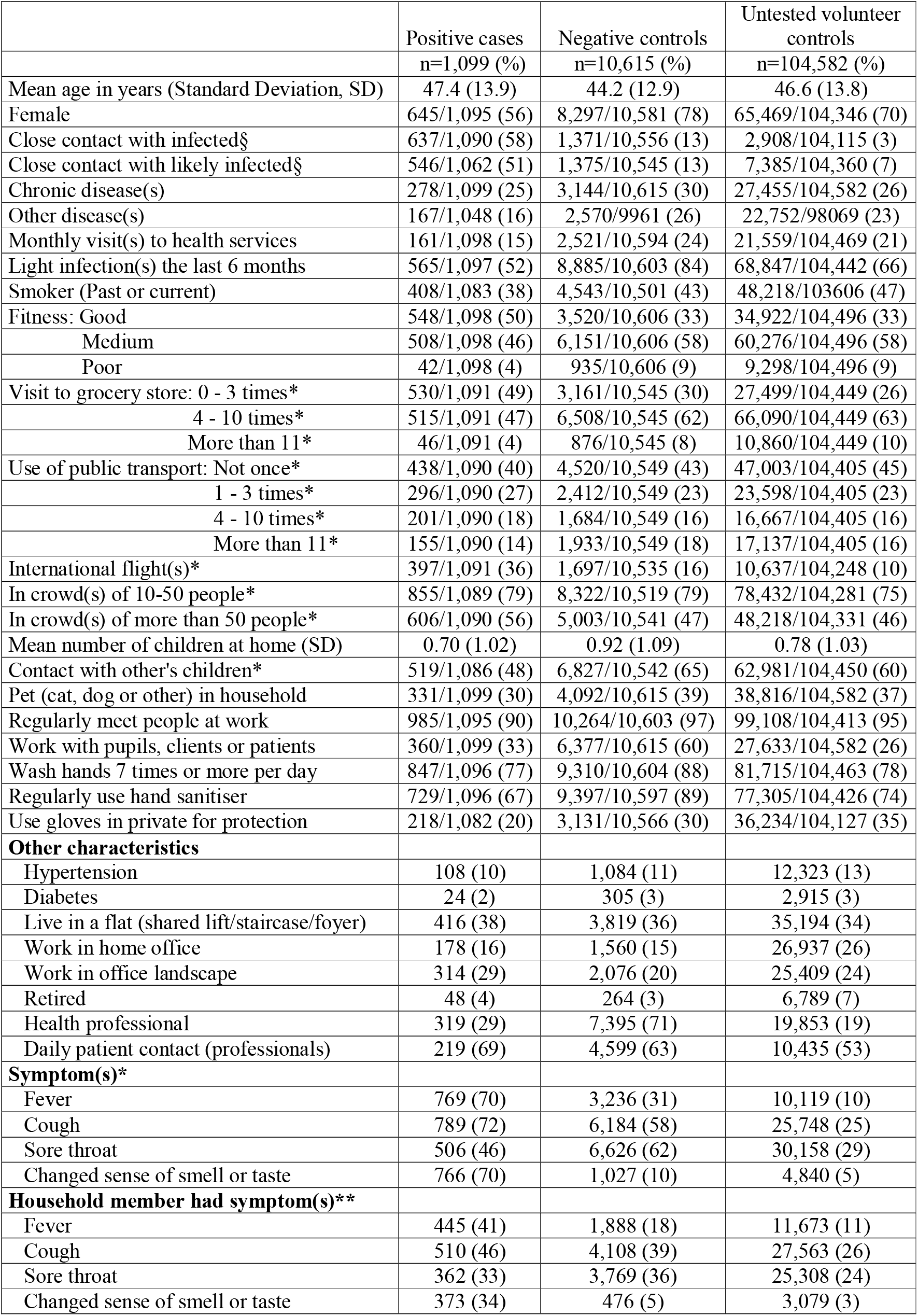
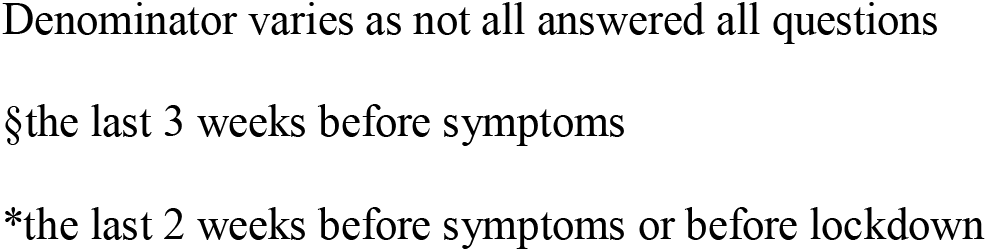
Study population characteristics

|  | Positive cases<br>n=1,099 (%) | Negative controls<br>n=10,615 (%) | Untested volunteer<br>controls<br>n=104,582 (%) |
| --- | --- | --- | --- |
| Mean age in years (Standard Deviation, SD) | 47.4 (13.9) | 44.2 (12.9) | 46.6 (13.8) |
| Female | 645/1,095 (56) | 8,297/10,581 (78) | 65,469/104,346 (70) |
| Close contact with infected§ | 637/1,090 (58) | 1,371/10,556 (13) | 2,908/104,115 (3) |
| Close contact with likely infected§ | 546/1,062 (51) | 1,375/10,545 (13) | 7,385/104,360 (7) |
| Chronic disease(s) | 278/1,099 (25) | 3,144/10,615 (30) | 27,455/104,582 (26) |
| Other disease(s) | 167/1,048 (16) | 2,570/9961 (26) | 22,752/98069 (23) |
| Monthly visit(s) to health services | 161/1,098 (15) | 2,521/10,594 (24) | 21,559/104,469 (21) |
| Light infection(s) the last 6 months | 565/1,097 (52) | 8,885/10,603 (84) | 68,847/104,442 (66) |
| Smoker (Past or current) | 408/1,083 (38) | 4,543/10,501 (43) | 48,218/103606 (47) |
| Fitness: Good | 548/1,098 (50) | 3,520/10,606 (33) | 34,922/104,496 (33) |
| Medium | 508/1,098 (46) | 6,151/10,606 (58) | 60,276/104,496 (58) |
| Poor | 42/1,098 (4) | 935/10,606 (9) | 9,298/104,496 (9) |
| Visit to grocery store: 0 - 3 times* | 530/1,091 (49) | 3,161/10,545 (30) | 27,499/104,449 (26) |
| 4 - 10 times* | 515/1,091 (47) | 6,508/10,545 (62) | 66,090/104,449 (63) |
| More than 11 * | 46/1,091 (4) | 876/10,545 (8) | 10,860/104,449 (10) |
| Use of public transport: Not once* | 438/1,090 (40) | 4,520/10,549 (43) | 47,003/104,405 (45) |
| 1 - 3 times* | 296/1,090 (27) | 2,412/10,549 (23) | 23,598/104,405 (23) |
| 4 - 10 times* | 201/1,090 (18) | 1,684/10,549 (16) | 16,667/104,405 (16) |
| More than 11 * | 155/1,090 (14) | 1,933/10,549 (18) | 17,137/104,405 (16) |
| International flight(s)* | 397/1,091 (36) | 1,697/10,535 (16) | 10,637/104,248 (10) |
| In crowd(s) of 10-50 people* | 855/1,089 (79) | 8,322/10,519 (79) | 78,432/104,281 (75) |
| In crowd(s) of more than 50 people* | 606/1,090 (56) | 5,003/10,541 (47) | 48,218/104,331 (46) |
| Mean number of children at home (SD) | 0.70 (1.02) | 0.92 (1.09) | 0.78 (1.03) |
| Contact with other's children* | 519/1,086 (48) | 6,827/10,542 (65) | 62,981/104,450 (60) |
| Pet (cat, dog or other) in household | 331/1,099 (30) | 4,092/10,615 (39) | 38,816/104,582 (37) |
| Regularly meet people at work | 985/1,095 (90) | 10,264/10,603 (97) | 99,108/104,413 (95) |
| Work with pupils, clients or patients | 360/1,099 (33) | 6,377/10,615 (60) | 27,633/104,582 (26) |
| Wash hands 7 times or more per day | 847/1,096 (77) | 9,310/10,604 (88) | 81,715/104,463 (78) |
| Regularly use hand sanitiser | 729/1,096 (67) | 9,397/10,597 (89) | 77,305/104,426 (74) |
| Use gloves in private for protection | 218/1,082 (20) | 3,131/10,566 (30) | 36,234/104,127 (35) |
| **Other characteristics** |  |  |  |
| Hypertension | 108 (10) | 1,084 (11) | 12,323 (13) |
| Diabetes | 24 (2) | 305 (3) | 2,915 (3) |
| Live in a flat (shared lift/staircase/foyer) | 416 (38) | 3,819 (36) | 35,194 (34) |
| Work in home office | 178 (16) | 1,560 (15) | 26,937 (26) |
| Work in office landscape | 314 (29) | 2,076 (20) | 25,409 (24) |
| Retired | 48 (4) | 264 (3) | 6,789 (7) |
| Health professional | 319 (29) | 7,395 (71) | 19,853 (19) |
| Daily patient contact (professionals) | 219 (69) | 4,599 (63) | 10,435 (53) |
| **Symptom(s)\*** |  |  |  |
| Fever | 769 (70) | 3,236 (31) | 10,119 (10) |
| Cough | 789 (72) | 6,184 (58) | 25,748 (25) |
| Sore throat | 506 (46) | 6,626 (62) | 30,158 (29) |
| Changed sense of smell or taste | 766 (70) | 1,027 (10) | 4,840 (5) |
| **Household member had symptom(s)\*\*** |  |  |  |
| Fever | 445 (41) | 1,888 (18) | 11,673 (11) |
| Cough | 510 (46) | 4,108 (39) | 27,563 (26) |
| Sore throat | 362 (33) | 3,769 (36) | 25,308 (24) |
| Changed sense of smell or taste | 373 (34) | 476 (5) | 3,079 (3) |

Denominator varies as not all answered all questions
§the last 3 weeks before symptoms
\*the last 2 weeks before symptoms or before lockdown

### Risk factors for SARS-COV-2 in 116.678 Norwegians

The final multivariable model included: age, sex, contact with infected individuals, chronic and other diseases, healthcare use, previous light infections, smoking, fitness, frequency of food shopping, public transport use, international flights, exposure to crowds, having children, contact with children, pets, meeting people at work, working with people, hand wash, hand sanitiser and use of gloves.

Table 2 compares cases with negative controls and untested volunteer controls. Having been on an international flight, and having had contact with a suspected or a confirmed COVID-19 case, was highly associated with SARS-CoV-2 positivity, in both control groups. Attending gatherings of more than 50 people was associated with a higher risk of SARS-CoV-2 positivity, whereas smaller gatherings were not. Those who used hand sanitiser or protective gloves privately had lower odds of SARS-CoV-2 positivity.

**Table 2.** Bivariate and multivariable analysis for SARS-CoV-2 risk factors.

|  | Bivariate | Multivariable |  | Bivariate | Multivariable |  |
| --- | --- | --- | --- | --- | --- | --- |
|  | Positive cases vs<br>Negative controls | Positive cases vs<br>Negative controls | P- | Positive cases vs<br>Untested volunteer<br>controls | Positive cases vs<br>Untested volunteer<br>controls | P- |
|  | OR (95% CI) | Adj. OR (95% CI) | value | OR (95% CI) | Adj. OR (95% CI) | value |
| Age in years | 1.02 (1.01 - 1.02) | 1.01 (1.00 - 1.01) | 0.123 | 1.00 (1.00 - 1.01) | 1.01 (1.00 - 1.02) | 0.001 |
| Male sex | 2.84 (2.49 - 3.22) | 1.52 (1.27 - 1.81) | <0.001 | 1.86 (1.65 - 2.09) | 1.51 (1.30 - 1.76) | <0.001 |
| Close contact with infected§ | 9.4 (8.3 - 10.8) | 4.56 (3.76 - 5.54) | <0.001 | 48.9 (43.2 - 55.5) | 24.4 (20.3 - 29.4) | <0.001 |
| Close contact with likely infected§ | 7.06 (6.18 - 8.06) | 3.13 (2.57 - 3.81) | <0.001 | 13.9 (12.3 - 15.7) | 2.80 (2.32 - 3.37) | <0.001 |
| Chronic disease(s) | 0.80 (0.70 - 0.93) | 0.94 (0.76 - 1.14) | 0.514 | 0.95 (0.83 - 1.09) | 1.21 (1.01 - 1.44) | 0.043 |
| Other disease(s) | 0.55 (0.46 - 0.65) | 0.72 (0.58 - 0.91) | 0.004 | 0.63 (0.53 - 0.74) | 0.78 (0.63 - 0.95) | 0.014 |
| Monthly visit(s) to health services | 0.55 (0.46 - 0.65) | 0.84 (0.66 - 1.07) | 0.154 | 0.66 (0.56 - 0.78) | 1.17 (0.94 - 1.45) | 0.162 |
| Light infection(s) the last 6 months | 0.21 (0.18 - 0.23) | 0.29 (0.24 - 0.34) | <0.001 | 0.55 (0.49 - 0.62) | 0.61 (0.53 - 0.71) | <0.001 |
| Smoker (Past or current) | 0.79 (0.70 - 0.90) | 0.87 (0.73 - 1.04) | 0.119 | 0.69 (0.61 - 0.79) | 0.82 (0.71 - 0.96) | 0.012 |
| Fitness: Good | 1 | 1 |  | 1 | 1 |  |
| Medium | 0.53 (0.47 - 0.60) | 0.78 (0.66 - 0.92) |  | 0.54 (0.48 - 0.61) | 0.72 (0.62 - 0.84) |  |
| Poor | 0.29 (0.21 - 0.40) | 0.49 (0.33 - 0.74) | <0.001 | 0.29 (0.21 - 0.39) | 0.48 (0.33 - 0.69) | <0.001 |
| Visit to grocery store: 0 - 3 times* | 1 | 1 |  | 1 | 1 |  |
| 4 - 10 times* | 0.47 (0.42 - 0.54) | 0.66 (0.55 - 0.78) |  | 0.40 (0.36 - 0.46) | 0.57 (0.49 - 0.66) |  |
| More than 11 * | 0.31 (0.23 - 0.43) | 0.46 (0.31 - 0.68) | <0.001 | 0.22 (0.16 - 0.30) | 0.32 (0.22 - 0.45) | <0.001 |
| Use of public transport: Not once* | 1 | 1 |  | 1 | 1 |  |
| 1 - 3 times* | 1.27 (1.08 - 1.48) | 1.15 (0.94 - 1.42) |  | 1.35 (1.16 - 1.56) | 1.15 (0.96 - 1.39) |  |
| 4 - 10 times* | 1.23 (1.03 - 1.47) | 1.01 (0.79 - 1.29) |  | 1.29 (1.09 - 1.53) | 1.02 (0.83 - 1.27) |  |
| More than 11* | 0.83 (0.68 - 1.00) | 0.93 (0.72 - 1.21) | 0.387 | 0.97 (0.81 - 1.17) | 0.91 (0.72 - 1.14) | 0.215 |
| International flight(s)* | 2.98 (2.61 - 3.41) | 1.85 (1.53 - 2.25) | <0.001 | 5.03 (4.44 - 5.70) | 3.60 (3.06 - 4.25) | <0.001 |
| In crowd(s) of 10-50 people* | 0.96 (0.83 - 1.12) | 0.89 (0.70 - 1.13) | 0.354 | 1.20 (1.04 - 1.39) | 1.00 (0.81 - 1.24) | 0.967 |
| In crowd(s) of more than 50 people* | 1.39 (1.22 - 1.57) | 1.21 (1.00 - 1.47) | 0.050 | 1.46 (1.29 - 1.64) | 1.32 (1.11 - 1.56) | 0.001 |
| Number of children | 0.81 (0.76 - 0.86) | 0.88 (0.81 - 0.96) | 0.003 | 0.92 (0.86 - 0.97) | 1.04 (0.96 - 1.12) | 0.338 |
| Contact with other's children* | 0.50 (0.44 - 0.56) | 0.77 (0.64 - 0.91) | 0.003 | 0.60 (0.53 - 0.68) | 0.71 (0.60 - 0.83) | <0.001 |
| Pet (cat, dog or other) in household | 0.69 (0.60 - 0.79) | 0.81 (0.68 - 0.97) | 0.021 | 0.73 (0.64 - 0.83) | 0.83 (0.71 - 0.97) | 0.022 |
| Regularly meet people at work | 0.30 (0.24 - 0.37) | 0.52 (0.37 - 0.73) | <0.001 | 0.48 (0.39 - 0.58) | 0.48 (0.36 - 0.62) | <0.001 |
| Work with pupils, clients or patients | 0.32 (0.28 - 0.37) | 0.54 (0.45 - 0.65) | <0.001 | 1.36 (1.20 - 1.54) | 1.24 (1.05 - 1.48) | 0.014 |
| Wash hands 7 times or more per day | 0.47 (0.41 - 0.55) | 0.92 (0.74 - 1.14) | 0.463 | 0.95 (0.82 - 1.09) | 1.10 (0.92 - 1.32) | 0.303 |
| Regularly use hand sanitiser | 0.25 (0.22 - 0.29) | 0.38 (0.31 - 0.46) | <0.001 | 0.70 (0.61 - 0.79) | 0.63 (0.54 - 0.75) | <0.001 |
| Use gloves in private for protection | 0.60 (0.51 - 0.70) | 0.54 (0.44 - 0.66) | <0.001 | 0.47 (0.41 - 0.55) | 0.55 (0.46 - 0.65) | <0.001 |

§the last three weeks before symptoms.
\*the last two weeks before symptoms or before lockdown.

Interestingly, having undergone “light” infections the last six months was strongly associated with lower odds ratio of SARS-CoV-2 positivity (Table 2). Contact with other people’s children or having children of your own (in preschool or in school), meeting multiple colleagues at work, and owning pets were also associated with a lower odds ratio of testing positive in comparison with both control groups.

The cases reported to have visited grocery stores fewer times than the controls the two weeks before a positive test. Unexpectedly, the cases reported to be more fit than the controls. This finding remained after the skiers who had been to Austria and Italy had been excluded from the analyses.

Table 2 shows that the two control groups yielded similar results, however, those who said they were working with clients, pupils or patients were found to have less SARS-CoV-2 compared with negative controls, but more SARS-CoV-2 than the untested. This effect disappeared in the analyses with health professionals only, whilst other aspects of the analyses remained unchanged (Table 3a).

**Table 3a.** Risk factors in health professionals

|  | Bivariate | Multivariable |  | Bivariate | Multivariable |  |
| --- | --- | --- | --- | --- | --- | --- |
|  | Cases vs Negative controls | Cases vs Negative controls | P- | Cases vs untested volunteer controls | Cases vs untested volunteer controls | P- |
|  | OR (95% CI) | Adj. OR (95% CI) | value | OR (95% CI) | Adj. OR (95% CI) | value |
| Age in years | 1.00 (0.99 - 1.01) | 1.00 (0.99 - 1.01) | 0.896 | 0.99 (0.98 - 1.00) | 1.01 (1.00 - 1.02) | 0.085 |
| Male sex | 1.49 (1.10 - 2.04) | 0.82 (0.56 - 1.22) | 0.323 | 1.64 (1.21 - 2.22) | 0.99 (0.68 - 1.43) | 0.946 |
| Close contact with infected§ | 7.04 (5.46 - 9.07) | 3.48 (2.44 - 4.98) | <0.001 | 13.1 (10.2 - 16.9) | 7.09 (4.76 - 10.57) | <0.001 |
| Close contact with likely infected§ | 6.43 (4.98 - 8.30) | 3.29 (2.30 - 4.72) | <0.001 | 8.27 (6.43 - 10.62) | 2.63 (1.76 - 3.92) | <0.001 |
| Chronic disease(s) | 0.69 (0.51 - 0.95) | 0.78 (0.53 - 1.14) | 0.193 | 0.84 (0.62 - 1.15) | 1.11 (0.77 - 1.61) | 0.581 |
| Other disease(s) | 0.92 (0.68 - 1.25) | 1.13 (0.78 - 1.64) | 0.511 | 1.00 (0.74 - 1.35) | 1.18 (0.83 - 1.68) | 0.350 |
| Monthly visit(s) to health services | 0.67 (0.46 - 0.96) | 0.80 (0.50 - 1.27) | 0.337 | 0.83 (0.58 - 1.19) | 1.02 (0.65 - 1.59) | 0.945 |
| Light infection(s) the last 6 months | 0.20 (0.16 - 0.26) | 0.24 (0.18 - 0.33) | <0.001 | 0.64 (0.50 - 0.82) | 0.71 (0.54 - 0.95) | 0.022 |
| Smoker (Past or current) | 0.77 (0.59 - 0.99) | 0.89 (0.65 - 1.23) | 0.491 | 0.74 (0.57 - 0.96) | 0.92 (0.68 - 1.24) | 0.572 |
| Fitness: Good | 1 | 1 |  | 1 | 1 |  |
| Medium | 0.57 (0.44 - 0.73) | 0.77 (0.57 - 1.04) |  | 0.62 (0.48 - 0.79) | 0.79 (0.59 - 1.05) |  |
| Poor | 0.11 (0.04 - 0.35) | 0.19 (0.06 - 0.61) | 0.001 | 0.15 (0.05 - 0.46) | 0.25 (0.08 - 0.81) | 0.010 |
| Visit to grocery store: 0 - 3 times* | 1 | 1 |  | 1 | 1 |  |
| 4 - 10 times* | 0.71 (0.55 - 0.93) | 0.77 (0.56 - 1.05) |  | 0.63 (0.48 - 0.81) | 0.69 (0.51 - 0.94) |  |
| More than 11 * | 0.46 (0.25 - 0.83) | 0.54 (0.28 - 1.06) | 0.106 | 0.36 (0.20 - 0.64) | 0.46 (0.25 - 0.88) | 0.013 |
| Use of public transport: Not once* | 1 | 1 |  | 1 | 1 |  |
| 1 - 3 times* | 1.71 (1.24 - 2.34) | 1.81 (1.23 - 2.65) |  | 2.05 (1.50 - 2.81) | 1.85 (1.29 - 2.66) |  |
| 4 - 10 times* | 1.72 (1.21 - 2.46) | 1.59 (1.02 - 2.47) |  | 2.46 (1.73 - 3.50) | 1.95 (1.29 - 2.94) |  |
| More than 11* | 1.37 (0.96 - 1.96) | 1.56 (1.00 - 2.45) | 0.017 | 2.12 (1.49 - 3.02) | 2.10 (1.38 - 3.20) | <0.001 |
| International flight(s)* | 2.13 (1.56 - 2.92) | 1.82 (1.23 - 2.70) | 0.004 | 3.23 (2.37 - 4.40) | 2.79 (1.91 - 4.07) | <0.001 |
| In crowd(s) of 10-50 people* | 0.73 (0.55 - 0.96) | 0.70 (0.47 - 1.03) | 0.075 | 0.93 (0.71 - 1.24) | 0.84 (0.58 - 1.21) | 0.353 |
| In crowd(s) of more than 50 people* | 0.95 (0.74 - 1.21) | 0.82 (0.58 - 1.14) | 0.239 | 1.13 (0.88 - 1.45) | 0.98 (0.71 - 1.34) | 0.888 |
| Number of children | 0.81 (0.72 - 0.92) | 0.88 (0.76 - 1.02) | 0.089 | 0.88 (0.78 - 0.99) | 1.07 (0.93 - 1.23) | 0.373 |
| Contact with other's children* | 0.57 (0.44 - 0.73) | 0.82 (0.60 - 1.12) | 0.207 | 0.62 (0.48 - 0.79) | 0.72 (0.53 - 0.97) | 0.030 |
| Pet (cat, dog or other) in household | 0.81 (0.62 - 1.05) | 0.98 (0.71 - 1.35) | 0.901 | 0.72 (0.56 - 0.93) | 0.83 (0.61 - 1.12) | 0.216 |
| Regularly meet people at work | 0.32 (0.14 - 0.70) | 0.30 (0.11 - 0.78) | 0.023 | 0.26 (0.12 - 0.57) | 0.32 (0.13 - 0.80) | 0.027 |
| Work with pupils, clients or patients | 1.16 (0.77 - 1.74) | 1.23 (0.75 - 2.03) | 0.399 | 1.34 (0.90 - 2.00) | 1.33 (0.82 - 2.14) | 0.230 |
| Wash hands 7 times or more per day | 0.50 (0.34 - 0.73) | 0.66 (0.42 - 1.05) | 0.088 | 0.56 (0.39 - 0.80) | 0.62 (0.40 - 0.97) | 0.045 |
| Regularly use hand sanitiser | 0.24 (0.16 - 0.36) | 0.16 (0.10 - 0.26) | <0.001 | 0.27 (0.19 - 0.39) | 0.20 (0.13 - 0.32) | <0.001 |
| Use gloves in private for protection | 0.48 (0.33 - 0.69) | 0.49 (0.31 - 0.76) | 0.001 | 0.49 (0.34 - 0.71) | 0.52 (0.34 - 0.80) | 0.001 |

§the last three weeks before symptoms.
\*the last two weeks before symptoms or before lockdown.

**Table 3b.** COVID-19 patients as a risk factors for health professionals

|  | Bivariate<br>Cases vs Negative<br>controls<br>(N=319 vs 7395) |  |  | Bivariate<br>Cases vs untested<br>volunteer controls<br>(N=319 vs 19853) |
| --- | --- | --- | --- | --- |
|  | OR (95% CI) |  |  | OR (95% CI) |
| Number of COVID-19 patients treated<br>in ward* |  |  |  |  |
| 0 | 1 (index) |  |  | 1 (index) |
| 1-5 | 1.2 (0.9-1.7) |  |  | 1.6 (1.2-2.3) |
| 6-10 | 1.8 (1.1-3.0) |  |  | 2.9 (1.7-4.8) |
| 11-20 | 1.2 (0.5-2.5) |  |  | 1.7 (0.8-3.6) |
| 21-50 | 1.3 (0.5-3.2) |  |  | 2.2 (0.9-5.5) |
| >51 | 1.8 (0.7-5.2) |  |  | 3.7 (1.3-10) |

\*Age and gender are included as confounders. Inclusion of all variables in the full model in Table 3a except “Close contact with infected” and “Close contact with likely infected” does not change the trend.

### Parents, teachers, child minders and pet owners

Having been in contact with other people’s children, or having children of your own, was associated with lower odds of SARS-CoV-2 positivity. In a separate analysis, parents of preschool children were found to have a lower odds of SARS-CoV-2 compared to others (Adj. OR 0.79, 95% CI 0.62-1.0), and likewise school children, even the older age groups, did not pose an increased risk for their parents (Adj. OR 0.83, 95% CI 0.69-0.99). The lack of risk in being a parent was also observed among health professionals. Neither having a cat (Adj. OR 0.84, 95% CI 0.67-1.07) nor a dog (Adj. OR 0.82, 95% CI 0.67-1.01) were risk factors for SARS-CoV-2.

### Risk factors for SARS-CoV-2 in health professionals

Practicing health professionals with patient contact constituted 18% (21,123) of the study population but 55 % of the tested individuals (7,642/11,608), p < 0.001), as per official testing policy (Supplement 2). Thirteen percent (2,722/21,054) of the tested health professionals suspected or knew that they had been exposed to a confirmed COVID-19 case compared to 7% (6,314/89,190) of the untested (OR 1.83, 95% CI 1.75-1.90). Furthermore, working in wards treating COVID-19 patients was associated with SARS-CoV-2 positivity both compared to SARS-CoV-2 negative and untested health professionals (Table 3b). There was no adverse effect in having children or having pets in this group. Among health professionals, there was a protective effect of washing hands more than seven times a day.

Use of public transport was significantly associated with SARS-CoV-2 positivity among health professionals (Table 3a). These effects remained unchanged when skiers (who had visited Austria/Italy) were removed.

### Sensitivity analyses

We performed sensitivity analyses where we excluded individuals who had been in close contact with infected, “skiers” (i.e. travellers to Austria and Italy), and health professionals (i.e. tested through the study period if they had symptoms) (Supplement 3). The above findings remained largely unchanged; the sample size was low (144 cases). Neither taking out the skiers, nor stratifying the controls on being formally invited versus recruited through mass media, changed the results (Supplement 3, Table 2).

## Discussion

This study indicates that being a parent, looking after children, and having pets were significantly associated with a lower risk of SARS-CoV-2 positivity. Furthermore, those who reported having undergone light infections in the last six months were less likely to test positive. The use of hand sanitiser was protective and health professionals who washed their hands more than seven times per day had significantly less SARS-CoV-2 infection. International travel, close contact with an infected person, and participation in large gatherings were highly associated with increased risk of SARS-CoV-2 positivity. Public transport risk was independent of dose (frequency) and was a risk factor for health professionals, likely because they have to travel in the rush hours.

The effect of children and pets on the risk of SARS-CoV-2 could be related to having been regularly exposed to low or non-pathogenic strains of the *Coronaviridae* family previously, possibly establishing some immunity to the disease through cross-reacting antibodies, mucosal immunity or enhanced unspecific viral immunity.^10^ Most light infections were likely upper respiratory tract viral infections, some of which might have been caused by members of the *Coronaviridae* family.^10^ We may speculate that such infections can induce resistance to SARS-CoV-2 infection, and/or ease COVID-19 disease.^10^ Having pets may induce changes to microbiome diversity and, likewise, induce immunological modulation, although research in this area is limited.^11^ The indicators of social connectedness were robustly and negatively associated with SARS-CoV-2 positivity. The possible protective effects of having undergone light infections, before the pandemic, is consistent with this hypothesis and, likewise, robustly confirmed in all analyses.

However, the “protection” received through contact with children could also have to do with behaviour that was not recorded in this study, such as parents may possibly socialise less with strangers, than non-parents.^12^ Furthermore, most of the positive participants were infected early in the pandemic in Norway. The virus was largely brought to the country by adults returning from ski-vacation in high prevalence areas and dissemination among children might occur at a later stage of the pandemic.

The main limitation of the study is that we tested fewer than 12.000 of the study population. Health professionals were tested widely throughout the study period. However, having COVID-19 symptoms was a pre-requisite for testing in all phases and we could only do a cross-sectional study. Due to the test policy, most of the negative controls were health professionals with other respiratory tract infections. Many other respiratory tract infections are transmitted in the same way as COVID-19. In terms of risk factors, the negative controls were therefore similar to the SARS-CoV-2 positive cases and some genuine risk factors probably did not reach significance in this comparison. We therefore included the untested group as a second control group of mostly healthy individuals. However, the difference between the untested and the background population was not determined and there are likely biases. Furthermore, less than 40% of the tested, invited individuals participated. Therefore, this study represents a conservative estimate of risk factors for SARS-CoV-2.

Old age and a number of clinical conditions associated with old age are risk factors for severe disease.^15^ This study recruited and asked questions on a digital platform. Although more than 69% of those aged 65-70 years in Norway use the internet on average one hour per day less than five percent of the study participants were above the age of 65 years.^16^ We cannot preclude that they represent those who were too ill to participate in a study. Similarly, very few of our participants were immigrants, refugees, or had been hospitalised, indicating that our data is skewed towards those less affected by SARS-CoV-2. Hard copy questionnaires or face-to-face interviews may be necessary to engage these sub-populations. Behavioural risk factors in this study may therefore not be applicable for those with severe disease, those who do not speak Norwegian, are unable to use digital media, and the oldest age group. Furthermore, study results cannot necessarily be extrapolated to other stages of the pandemic. However, for those who were included, the comprehensive questionnaire was fully completed by the vast majority of the participants and most submitted their responses less than three weeks after lockdown, indicating that information on pre-lockdown behaviour may be reliable and that the study describes viral dissemination in a susceptible population almost completely without any societal countermeasures.

Having had contact with a likely or a confirmed case of COVID-19 posed the highest risk of acquiring SARS-CoV-2. Restrictions on large gatherings and all gatherings that may involve contact with infected as well as restrictive travel recommendations seem warranted. Based on our results, we recommend continued injunction on large gatherings during the pandemic. Furthermore, well-placed and free hand sanitizer in public places and persistent cleaning with soap and water where hands may touch surfaces may be relatively inexpensive and probably very efficient in preventing SARS-CoV-2 infection. Importantly, further research is needed to confirm whether children and social interaction can induce resistance to SARS-CoV-2 in other settings and in the next phases of the pandemic.

## Supporting information

Statistical analysis plan

STROBE checklist

## Data Availability

Data sharing is restricted by the informed consent form, the ethical approval committee and the European General Data Protection Regulation (GDPR). Upon request, in agreement with the above, the PI may be able grant partial access to an anonymized data set.

## Acknowledgements

We thank Dagfinn Bergsager, Stein-Erik Lund, Milen Kouylekov, Olaug Reiakvam, and Roy Farai Manyaira for their help with data collection, manuscript preparation or submission. We thank the Age Labs AS for allowing AS, KTK and CLH to work on this project.

## Data sharing

Date sharing is restricted by the informed consent form, the ethical approval committee and the European General Data Protection Regulation (GDPR). Upon request, in agreement with the above, the PI may be able grant partial access to an anonymized data set.

## Contributors

AS, CLS, JAD and KTK designed the study.

AS, CLS, CLH, SJ, FOP, BH, JAD, MI and KTK designed the questionnaire.

AS, KTK, LBS, EA, AL, RB, EFK, SBJ and MI recruited participants and administered the data collection.

TÅM, EFK, AS, GU, KL and KTK performed data cleaning, statistical analysis and interpretation.

EFK, AS, KTK, GU, ER, BH, MI, CLH, CLS, SJ, FOP, EA, LBS, RB, SBJ, KL, JAD and TÅM wrote the article. All authors reviewed and approved the article before submission.

## Supplementary Material

### Supplement 1 – Comparison of tested Invited and Volunteers

#### Differences Between Tested Invited Cases and Controls and Volunteers with Self-reported Test Results

A total of 7839 participants were included after they were invited to the study because of a SARS-CoV-2 PCR test (“Invited”). Another 108839 participants volunteered to participate after media coverage of the study. 3875 of these participants self-reported a SARS-CoV-2 PCR test result.

In Table S1a the laboratory confirmed invited cases and controls were compared to the group of volunteers that self-reported a test result. The table shows that the groups were broadly similar. The volunteers used less public transportation and more often lived in a free-standing hose than the Invited. The invited were recruited from a relatively urbanized area in Norway whereas the Volunteers came from the whole country including more rural areas.

**Table S1a.**
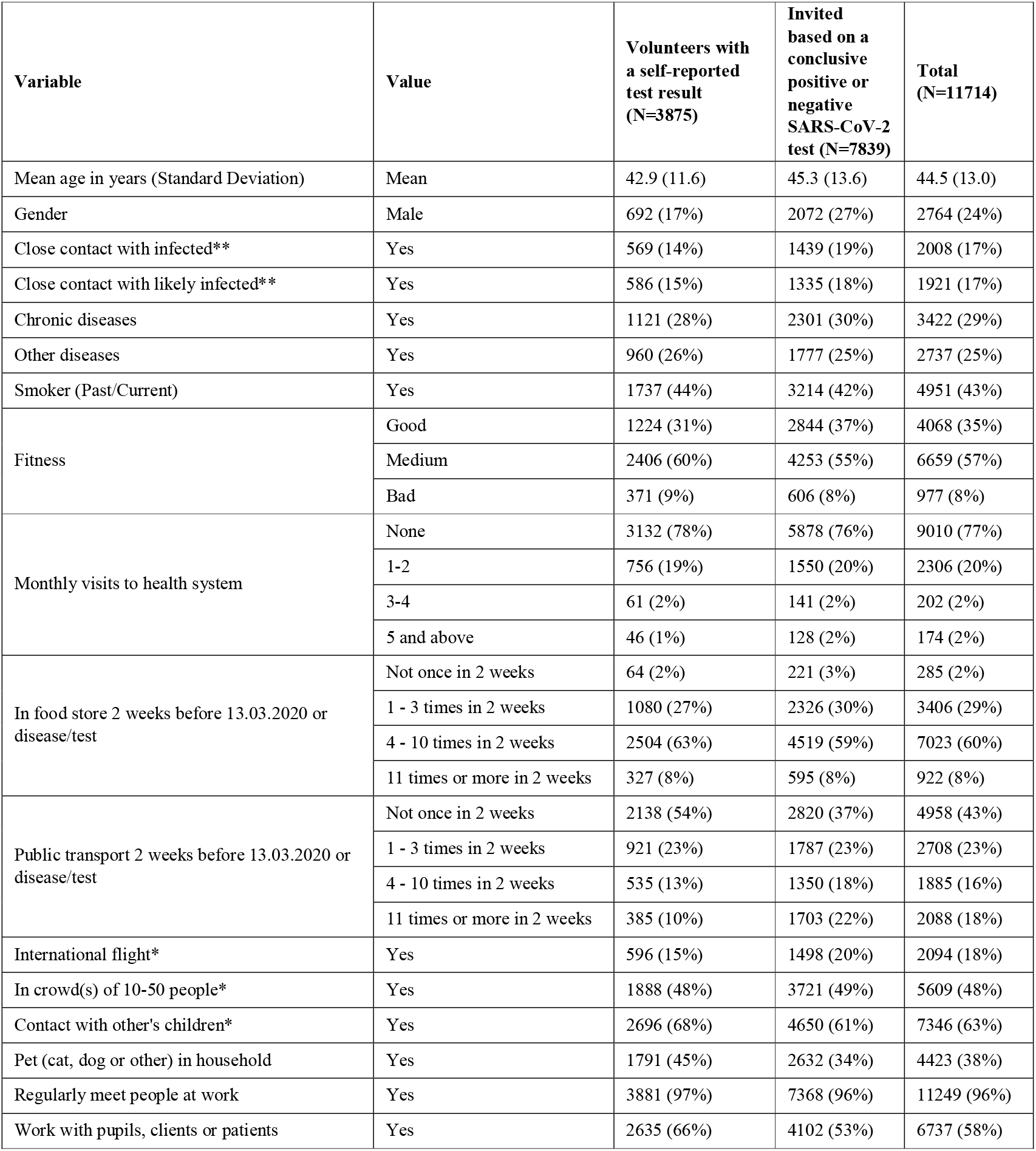
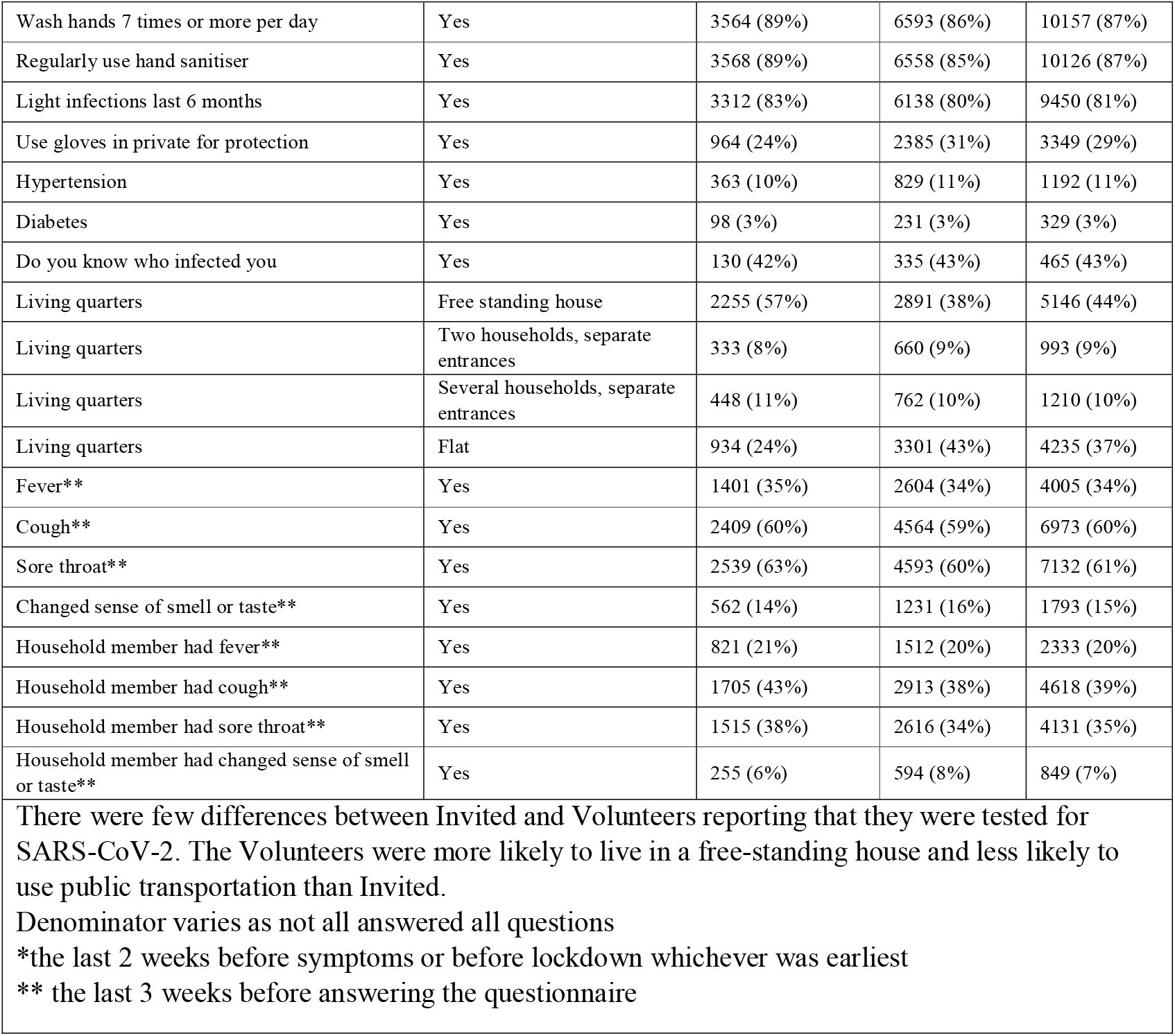

| Variable | Value | Volunteers with a self-reported test result (N=3875) | Invited based on a conclusive positive or negative SARS-CoV-2 test (N=7839) | Total (N=11714) |
| --- | --- | --- | --- | --- |
| Mean age in years (Standard Deviation) | Mean | 42.9 (11.6) | 45.3 (13.6) | 44.5 (13.0) |
| Gender | Male | 692 (17%) | 2072 (27%) | 2764 (24%) |
| Close contact with infected** | Yes | 569 (14%) | 1439 (19%) | 2008 (17%) |
| Close contact with likely infected** | Yes | 586 (15%) | 1335 (18%) | 1921 (17%) |
| Chronic diseases | Yes | 1121 (28%) | 2301 (30%) | 3422 (29%) |
| Other diseases | Yes | 960 (26%) | 1777 (25%) | 2737 (25%) |
| Smoker (Past/Current) | Yes | 1737 (44%) | 3214 (42%) | 4951 (43%) |
| Fitness | Good | 1224 (31%) | 2844 (37%) | 4068 (35%) |
|  | Medium | 2406 (60%) | 4253 (55%) | 6659 (57%) |
|  | Bad | 371 (9%) | 606 (8%) | 977 (8%) |
| Monthly visits to health system | None | 3132 (78%) | 5878 (76%) | 9010 (77%) |
|  | 1-2 | 756 (19%) | 1550 (20%) | 2306 (20%) |
|  | 3-4 | 61 (2%) | 141 (2%) | 202 (2%) |
|  | 5 and above | 46 (1%) | 128 (2%) | 174 (2%) |
| In food store 2 weeks before 13.03.2020 or disease/test | Not once in 2 weeks | 64 (2%) | 221 (3%) | 285 (2%) |
|  | 1 - 3 times in 2 weeks | 1080 (27%) | 2326 (30%) | 3406 (29%) |
|  | 4 - 10 times in 2 weeks | 2504 (63%) | 4519 (59%) | 7023 (60%) |
|  | 11 times or more in 2 weeks | 327 (8%) | 595 (8%) | 922 (8%) |
| Public transport 2 weeks before 13.03.2020 or disease/test | Not once in 2 weeks | 2138 (54%) | 2820 (37%) | 4958 (43%) |
|  | 1 - 3 times in 2 weeks | 921 (23%) | 1787 (23%) | 2708 (23%) |
|  | 4 - 10 times in 2 weeks | 535 (13%) | 1350 (18%) | 1885 (16%) |
|  | 11 times or more in 2 weeks | 385 (10%) | 1703 (22%) | 2088 (18%) |
| International flight* | Yes | 596 (15%) | 1498 (20%) | 2094 (18%) |
| In crowd(s) of 10-50 people* | Yes | 1888 (48%) | 3721 (49%) | 5609 (48%) |
| Contact with other's children* | Yes | 2696 (68%) | 4650 (61%) | 7346 (63%) |
| Pet (cat, dog or other) in household | Yes | 1791 (45%) | 2632 (34%) | 4423 (38%) |
| Regularly meet people at work | Yes | 3881 (97%) | 7368 (96%) | 11249 (96%) |
| Work with pupils, clients or patients | Yes | 2635 (66%) | 4102 (53%) | 6737 (58%) |
| Wash hands 7 times or more per day | Yes | 3564 (89%) | 6593 (86%) | 10157 (87%) |
| Regularly use hand sanitiser | Yes | 3568 (89%) | 6558 (85%) | 10126 (87%) |
| Light infections last 6 months | Yes | 3312 (83%) | 6138 (80%) | 9450 (81%) |
| Use gloves in private for protection | Yes | 964 (24%) | 2385 (31%) | 3349 (29%) |
| Hypertension | Yes | 363 (10%) | 829 (11%) | 1192 (11%) |
| Diabetes | Yes | 98 (3%) | 231 (3%) | 329 (3%) |
| Do you know who infected you | Yes | 130 (42%) | 335 (43%) | 465 (43%) |
| Living quarters | Free standing house | 2255 (57%) | 2891 (38%) | 5146 (44%) |
| Living quarters | Two households, separate entrances | 333 (8%) | 660 (9%) | 993 (9%) |
| Living quarters | Several households, separate entrances | 448 (11%) | 762 (10%) | 1210 (10%) |
| Living quarters | Flat | 934 (24%) | 3301 (43%) | 4235 (37%) |
| Fever** | Yes | 1401 (35%) | 2604 (34%) | 4005 (34%) |
| Cough** | Yes | 2409 (60%) | 4564 (59%) | 6973 (60%) |
| Sore throat** | Yes | 2539 (63%) | 4593 (60%) | 7132 (61%) |
| Changed sense of smell or taste** | Yes | 562 (14%) | 1231 (16%) | 1793 (15%) |
| Household member had fever** | Yes | 821 (21%) | 1512 (20%) | 2333 (20%) |
| Household member had cough** | Yes | 1705 (43%) | 2913 (38%) | 4618 (39%) |
| Household member had sore throat** | Yes | 1515 (38%) | 2616 (34%) | 4131 (35%) |
| Household member had changed sense of smell or taste** | Yes | 255 (6%) | 594 (8%) | 849 (7%) |
| There were few differences between Invited and Volunteers reporting that they were tested for SARS-CoV-2. The Volunteers were more likely to live in a free-standing house and less likely to use public transportation than Invited.<br>Denominator varies as not all answered all questions<br>*the last 2 weeks before symptoms or before lockdown whichever was earliest<br>** the last 3 weeks before answering the questionnaire |  |  |  |  |

#### Correspondence Between Self-reported and Laboratory Confirmed SARS-CoV-2 PCR Tests

**Table S1b.**

|  |  | Laboratory confirmed SARS-CoV-2 PCR test result |  |  |  |
| --- | --- | --- | --- | --- | --- |
|  |  | Negative for SARS-CoV-2 PCR | Positive for SARS-CoV-2 PCR | Unknown at the time of data collection from lab* | Total |
| **Self-reported test result** | Negative for SARS-CoV-2 PCR | 6881 | 0 | 45 | 6926 |
|  | Positive for SARS-CoV-2 PCR | 11 | 754 | 21 | 786 |
|  | Waiting for result | 141 | 7 | 42 | 190 |
|  | Not tested | 58 | 5 | 23 | 86 |
|  | Total | 7091 | 766 | 131 | 7988 |
| Very good correspondence between self-reported test result and laboratory confirmed test results in invited participants (kappa 0.99).<br>* Including inconclusive test results |  |  |  |  |  |

To confirm that the Volunteers reported their SARS-CoV-2 PCR test results faithfully, laboratory results from Volunteers already included in the study were obtained from a nearby lab that was not a source of Invited participants. The table shows that these Volunteers had self-reported their test results precisely (kappa 0.97).

**Table S1c.**

|  |  | Laboratory confirmed SARS-CoV-2 PCR test result for Volunteer group obtained after data collection. |  |  |  |
| --- | --- | --- | --- | --- | --- |
|  |  | Negative for SARS-CoV-2 PCR | Positive for SARS-CoV-2 PCR | Unknown at the time of data collection from lab* | Total |
| **Self-reported test result from Volunteer group** | Negative for SARS-CoV-2 PCR | 465 | 0 | 1 | 466 |
|  | Positive for SARS-CoV-2 PCR | 2 | 34 | 3 | 39 |
|  | Waiting for result | 2 | 0 | 1 | 3 |
|  | Not tested | 0 | 0 | 0 | 0 |
|  | Total | 469 | 34 | 5 | 508 |
| Very good correspondence between self-reported test result and laboratory results in Volunteer participants tested at one particular lab (kappa 0.97).<br>* Including inconclusive test results. |  |  |  |  |  |

### Supplement 2 – Healthcare Professionals with Patient Contact

Healthcare professionals with and without patient constituted 24% of the study population and healthcare professionals with patient contact at least weekly constituted 18% of the study population. The latter group were tested for SARS-CoV-2 if they had symptoms throughout the study period and was therefore analyzed separately in the paper. In Table S2 healthcare professionals with patient contact are compared with the rest of the study population. Men were underrepresented among healthcare professionals with patient contact.

**Table S2.**

| Variable | Value | All participants without patient contact<br>(N=95555) | Healthcare professionals with patient contact#<br>(N=21123) | Total<br>(N=116678) |
| --- | --- | --- | --- | --- |
| Mean age in years (Standard Deviation) | Median | 46-50 years | 41-45 years | 41-45 years |
| Gender | Male | 30815 (32%) | 2915 (14%) | 33730 (29%) |
| SARS-CoV-2 positive | Yes | 837 (0.9%) | 262 (1.2%) | 1099 (0.9%) |
| Close contact with infected** | Yes | 2851 (3%) | 2135 (10%) | 4986 (4%) |
| Close contact with likely infected** | Yes | 6667 (7%) | 2722 (13%) | 9389 (8%) |
| Chronic diseases | Yes | 26275 (28%) | 4734 (22%) | 31009 (27%) |
| Other diseases | Yes | 21205 (24%) | 4391 (22%) | 25596 (23%) |
| Smoker (Past/Current) | Yes | 44625 (47%) | 8717 (42%) | 53342 (46%) |
| Fitness | Very fit | 31521 (33%) | 7584 (36%) | 39105 (34%) |
|  | Fairly fit | 54909 (58%) | 12254 (58%) | 67163 (58%) |
|  | In bad shape | 9037 (10%) | 1277 (6%) | 10314 (9%) |
| Monthly visits to health system | None | 74511 (78%) | 17685 (84%) | 92196 (79%) |
|  | 43862 | 18359 (19%) | 3069 (15%) | 21428 (18%) |
|  | 43924 | 1592 (2%) | 180 (1%) | 1772 (2%) |
|  | 5 and above | 992 (1%) | 155 (1%) | 1147 (1%) |
| In food store 2 weeks before 13.03.2020 or disease/test* | Not once in 2 weeks | 1553 (2%) | 175 (1%) | 1728 (2%) |
|  | 1 - 3 times in 2 weeks | 24518 (26%) | 5070 (24%) | 29588 (25%) |
|  | 4 - 10 times in 2 weeks | 59431 (62%) | 13898 (66%) | 73329 (63%) |
|  | 11 times or more in 2 weeks | 9895 (10%) | 1927 (9%) | 11822 (10%) |
| Public transport 2 weeks before 13.03.2020 or disease/test* | Not once in 2 weeks | 41732 (44%) | 10381 (49%) | 52113 (45%) |
|  | 1 - 3 times in 2 weeks | 21761 (23%) | 4624 (22%) | 26385 (23%) |
|  | 4 - 10 times in 2 weeks | 15853 (17%) | 2779 (13%) | 18632 (16%) |
|  | 11 times or more in 2 weeks | 16009 (17%) | 3287 (16%) | 19296 (17%) |
| International flight* | Yes | 11042 (12%) | 1746 (8%) | 12788 (11%) |
| In crowd(s) of 10-50 people* | Yes | 45092 (47%) | 8920 (42%) | 54012 (46%) |
| Contact with other's children* | Yes | 56461 (59%) | 14113 (67%) | 70574 (61%) |
| Pet | Yes | 34764 (36%) | 8611 (41%) | 43375 (37%) |
| Regularly meet people at work | Yes | 89795 (94%) | 20934 (99%) | 110729 (95%) |
| Job with people (pupils, clients, patients) | Yes | 16159 (17%) | 18415 (87%) | 34574 (30%) |
| Wash hands 7 times or more per day | Yes | 72675 (76%) | 19523 (93%) | 92198 (79%) |
| Regularly use hand sanitizer | Yes | 67431 (71%) | 20302 (96%) | 87733 (75%) |
| Light infections last 6 months | Yes | 63456 (67%) | 15124 (72%) | 78580 (67%) |
| Use gloves in private for protection | Yes | 34750 (37%) | 4941 (24%) | 39691 (34%) |
| Hypertension | Yes | 11835 (13%) | 1680 (8%) | 13515 (12%) |
| Diabetes | Yes | 2814 (3%) | 430 (2%) | 3244 (3%) |
| Do you know who infected you? | Yes | 350 (42%) | 115 (44%) | 465 (43%) |
| Living quarters | Free standing house | 44250 (47%) | 10093 (48%) | 54343 (47%) |
|  | Two households, separate entrances | 8008 (9%) | 1758 (8%) | 9766 (9%) |
|  | Several households, separate entrances | 9704 (10%) | 2093 (10%) | 11797 (10%) |
|  | Flat | 32454 (34%) | 6975 (33%) | 39429 (34%) |
| Fever** | Yes | 11507 (12%) | 2617 (12%) | 14124 (12%) |
| Cough** | Yes | 26647 (28%) | 6074 (29%) | 32721 (28%) |
| Sore throat** | Yes | 29782 (31%) | 7508 (36%) | 37290 (32%) |
| Changed sense of smell or taste** | Yes | 5556 (6%) | 1077 (5%) | 6633 (6%) |
| Household member had fever** | Yes | 11668 (12%) | 2338 (11%) | 14006 (12%) |
| Household member had cough** | Yes | 26344 (28%) | 5837 (28%) | 32181 (28%) |
| Household member had sore throat** | Yes | 23943 (25%) | 5496 (26%) | 29439 (25%) |
| Household member had changed sense of smell or taste** | Yes | 3361 (4%) | 567 (3%) | 3928 (3%) |
| Comparison between study participants without direct patient contact and healthcare professionals with patient contact at least once per week.<br>Denominator varies as not all answered all questions<br>*the last 2 weeks before symptoms or before lockdown whichever came first<br>** the last 3 weeks before answering the questionnaire<br># Healthcare professionals with patient contact at least once per week |  |  |  |  |

### Supplement 3 – Sensitivity Analysis

#### Sensitivity analysis without high-risk individuals

Participants with a travel history to Italy or Austria and reporting contact with a person with likely or confirmed SARS-CoV-2 infection as well as health professionals working in clinic were excluded from this analysis because they had a likely known source of their infection. The number of remaining cases were 144.

**Table S3-1.**

| Variable | Level/Category | Multivariate analysis<br>Positive cases vs<br>Negative controls (OR,<br>95% CI)<br>(N=3690) | Multivariate analysis<br>Positive cases vs<br>Untested<br>volunteers controls<br>(OR, 95% CI)<br>(N=83486) |
| --- | --- | --- | --- |
| Gender | Male | 1.73 (1.16 – 2.58) | 1.94 (1.33 – 2.83) |
| Chronic diseases | Yes | 1.33 (0.86 – 2.07) | 1.66 (1.10 – 2.50) |
| Other diseases | Yes | 0.49 (0.28 – 0.83) | 0.65 (0.39 – 1.09) |
| Smoking | Past/Current | 0.69 (0.46 – 1.04) | 0.73 (0.50 – 1.07) |
| Fitness | Good | Reference. | Reference |
|  | Medium | 0.99 (0.66 – 1.49) | 0.94 (0.64 – 1.37) |
|  | Bad | 0.55 (0.22 – 1.35) | 0.55 (0.23 – 1.31) |
| Prior visits in health service | Yes | 0.61 (0.36 – 1.04) | 1.00 (0.60 – 1.65) |
| Visits to food store last 2 weeks* | 0-3 | Reference. | Reference |
|  | 4-10 | 0.62 (0.42 – 0.92) | 0.46 (0.31 – 0.66) |
|  | 11+ | 0.22 (0.07 – 0.73) | 0.14 (0.04 – 0.44) |
| Use of public transportation past 2 *weeks | 0 | Reference. | Reference |
|  | 1-3 | 0.73 (0.45 – 1.19) | 0.86 (0.54 – 1.37) |
|  | 4-10 | 0.72 (0.41 – 1.26) | 0.88 (0.51 – 1.52) |
|  | 11+ | 0.66 (0.36 – 1.23) | 0.86 (0.48 – 1.55) |
| International flights past 2 weeks* | Yes | 0.89 (0.57 – 1.39) | 2.71 (1.78 – 4.11) |
| Being in crowds of 10-50 people past 2 weeks* | Yes | 1.31 (0.77 – 2.21) | 1.30 (0.80 – 2.14) |
| Being in crowds of 50+ people last 2 weeks* | Yes | 0.98 (0.63 – 1.53) | 0.95 (0.63 – 1.43) |
| Contact with other kids past 2 weeks* | Yes | 0.80 (0.53 – 1.20) | 0.76 (0.51 – 1.11) |
| Pet | Yes | 0.68 (0.45 – 1.03) | 0.78 (0.52 – 1.15) |
| Regularly meet people at work | Yes/NA | 0.78 (0.38 – 1.60) | 0.70 (0.36 – 1.38) |
| Work with pupils, clients or patients | Yes | 1.49 (0.93 – 2.39) | 2.08 (1.33 – 3.23) |
| Wash hands 7 times or more per day | 7+ | 1.03 (0.65 – 1.63) | 1.32 (0.86 – 2.02) |
| Regularly use hand sanitiser | Yes | 0.57 (0.38 – 0.86) | 0.88 (0.60 – 1.29) |
| Light infections last 6 months | Yes | 0.26 (0.18 – 0.38) | 0.61 (0.43 – 0.89) |
| Use gloves in private for protection | Yes | 0.44 (0.27 – 0.70) | 0.45 (0.29 – 0.71) |
| Number of children at home | Continuous | 0.91 (0.75 – 1.11) | 1.04 (0.86 – 1.26) |
| Age | Continuous | 1.00 (0.99 – 1.02) | 1.00 (0.99 – 1.02) |
| The main findings in the study remained robust in multivariate analysis where high-risk individuals were omitted from the case group. However, the confidence intervals are wide because of the low number of cases included in this analyses.<br>CI: confidence interval, OR: odds ratio<br>*the last 2 weeks before symptoms or before lockdown whichever was earliest |  |  |  |

#### Sensitivity Analysis without Self-reported SARS-CoV-2 PCR Positive Cases

The study was planned as an unmatched case-control study among all patients tested positively and negatively for SARS-CoV-2 in several large laboratories as the cases and controls, respectively. An additional control group of untested volunteers were recruited by self referral. In total, 759 Cases with PCR confirmed SARS-CoV-2, 7080 Controls with a negative SARS-CoV-2 result and 104.964 Volunteer Controls reporting not to have undergone testing for SARS-CoV-2 were recruited. The findings in the Case-Control study were fully in accordance with the somewhat larger cross-sectional study.

**Table S3-2.**
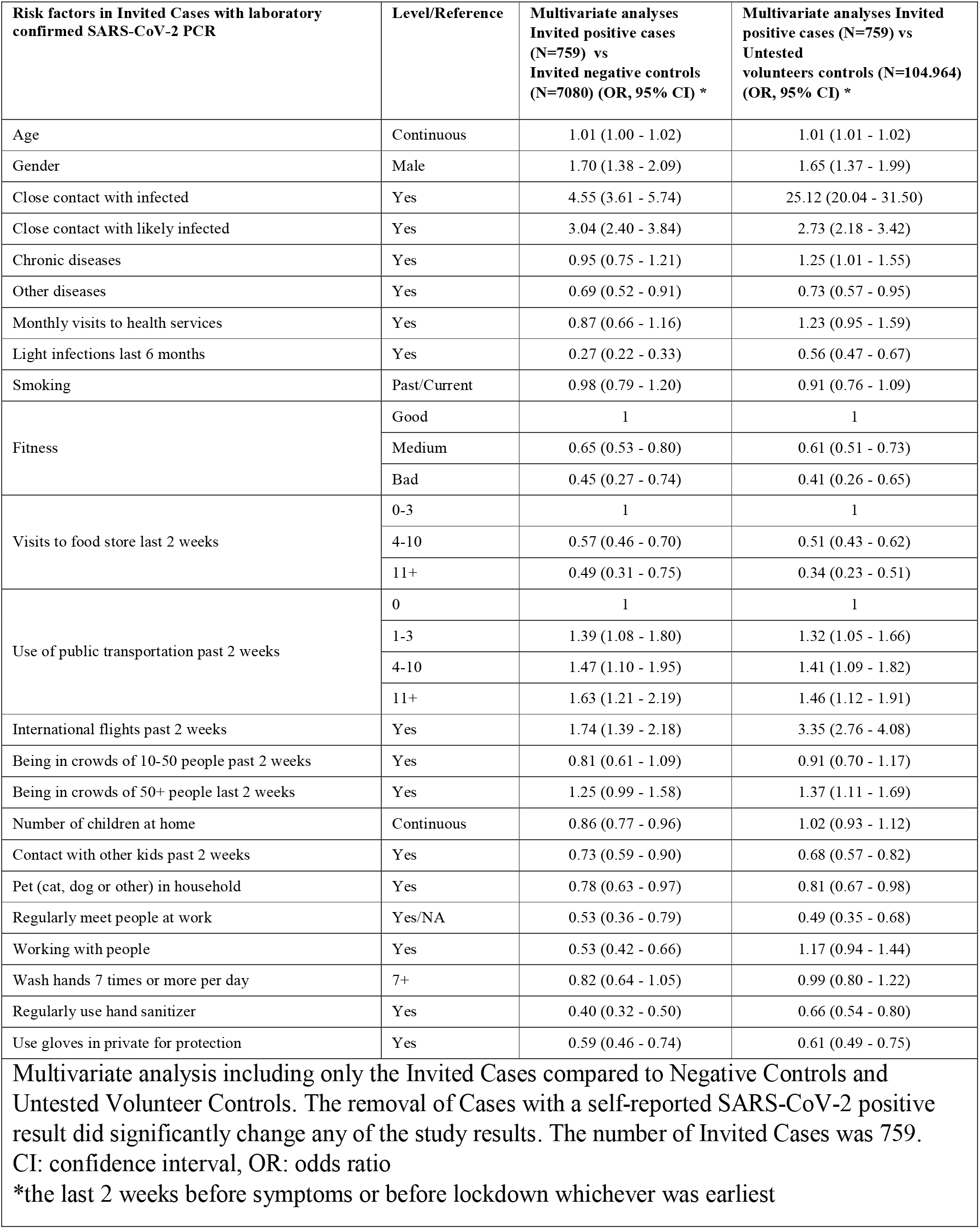

