## Supplementary material for "Risk factors for community transmission of SARS-CoV-2. A cross-sectional study in 116,678 people": Statistical analysis plan

### Risk factors for community- and workplace transmission of COVID-19

v. 08 2020

#### Statistical analysis plan

The aim of the study is to identify risk factors associated with acquisition of COVID-19 virus. The study is designed as an unmatched case-control study, as well as a cross-sectional study. The study can be developed into a cohort study with future data collections. Analysis of that part is not covered in this plan.

##### Study population

*Invited*

Participants were identified from testing laboratories at Oslo University Hospital, Vestre Viken and one private laboratory (Fürst). Combined these three laboratories cover an estimated 70% of the population in and around Oslo. The study population included all individuals who had been tested at these three laboratories during the period January 2020-April 6. All individuals who had tested positive were defined as "cases" and those who had tested negative were defined as "controls". All cases and controls were invited (March 27-April 4) to participate in a study entitled "Risk factors for becoming infected with coronavirus at work or in the community". The invitations were sent as direct text messaging (SMS) to the cell phone numbers registered where they were asked to log in with their unique Norwegian personal ID number, read and sign an informed consent and answer a questionnaire.

*Volunteers*

A general message on an ongoing study with the same title was publicised through media on March 27 and was covered by national media, including TV on March 29th. Volunteers were encouraged to participate by completing the same online questionnaire as those invited. Participants were asked to log in with their unique Norwegian personal ID number, read and sign an informed consent and answer the questionnaire.

Differences in sampling methods and the data collected give possibilities to define different study populations:

1. Study population P1:
    a. Cases -> Positive SARS-CoV-19 result reported from laboratory
    b. Controls 1-> Negative SARS-CoV-19 result reported from laboratory

    c. Controls 2 -> Untested population of volunteers
2. Study population P2:
    a. Cases -> Reported a positive test in the questionnaire
    b. Controls 1 -> Reported a negative test in the questionnaire
    c. Controls 2 -> Untested population of volunteers

##### Testing of COVID-19 in the study population

WHO recommends testing every person that has a fever and at least one symptom of COVID-19. Testing capacity is limited in Norway and the National Institute of Public Health (NIPH) therefore has introduced criteria to limit testing.

The most important point about these criteria is that until March 12, testing was open to anybody with COVID-19 symptoms a relatively high proportion of infections were detected (defined as “early” in the study). However, after the change in guidelines was made March 12 only prioritized groups were tested (defined as “late” in the study).

History of changes in testing rules in Norway

- From January: All suspected cases. I.e: Symptoms AND travel history. The suspected cases criteria were changed gradually to cover all suspected cases during the epidemic.
- February 25: Hospitalized patients with unexplained viral pneumonia added to test group
- March 8: Healthcare personal with any symptoms from the airways within 14 days of traveling abroad added to the test group.
- March 10: All hospitalized patients with acute respiratory symptoms added to the test group.
- March 12: **End of testing of any suspected case was announced this day.** From now on only healthcare personnel with patient contact and airway symptoms, close contacts of a confirmed case IF contact tracing is indicated, hospitalized patients and vulnerable groups (>65 yo+cardiovascular, lung, cancer, hypertension or diabetics) with minor airway symptoms were tested

- April 5: If good capacity in testing it is recommended to test anybody with COVID-19 symptoms.

#### Analyses

A descriptive table comparing summary statistics or distributions for demographic variables and potential risk factors, stratified on cases and controls will be made - separately for P1 and P2.

Univariate and multivariate logistic regressions will be estimated for all study populations. The outcome is case/control (cf. definitions of study populations above), and the independent variables are a priori defined confounders and a list of potential and relevant risk factors. Stratified analyses across sex, age, working group (people working in health sector vs others) and early testers vs later testers (after March 12) will be done to assess robustness of results.

Due to limited access to information on non-responders, simple comparisons on distribution of age and sex between responders and non-responders will be made to assess generalisability. These analyses will be stratified by case/controls.

#### Confounding and simplification strategy

As a starting point, a list of a priori potential risk factor are made (see list below). In a study like this, with many prognostic factors, the variables are likely to confound each other. We consider the demographic variables sex and age to be important (mandatory) confounders, and these factors will thus be included in all multivariate regressions. To ease presentation and convey important findings the number of independent variables should be reduced. We will do this by removing those variables that are either evenly distributed among case/controls, or have an effect estimate very close to 1 in multivariable regressions AND that does not alter estimated effects on the other variables in the regression models.

#### Potential risk factors

The questionnaire provides information on a number of different variables. Below follows a summary and description of the selected confounders and risk factors, listed (almost) in the order as they occur in the questionnaire:

1. Anxiety for Covid-19
    a. Yes = "Veldig"
    b. No = "Nei/Vet ikke"
2. Close contact with others likely to be infected with Covid-19
    a. Yes = "Nærkontakt med sikkert smittet eller med sannsynlig smittet"
3. Working in the health sector
    a. Yes = "Ja"
    b. No = "Nei/Vet ikke"
4. Morbidity/chronic diseases
    a. Yes = At least one of the conditions listed: Heart disease, hypertension, chronic lung disease, asthma, diabetes, immune-supressed, cancer
5. Obesity
    a. Bmi >= 30
6. Smoking
    a. Yes = "Ja/tidligere"
7. Physical fitness
    a. Bad = "Dårlig"
    b. Medium = "Middels"
    c. Good = "God"
8. Shopping habits
    a. Not often = "Vært i matbutikk 3 eller færre ganger"
    b. Frequent = "Vært i matbutikk 4 eller flere ganger"
9. Commuting
    a. Not often = "Reist kollektivt 3 eller færre ganger"
    b. Frequent = "Reist kollektivt 4 eller flere ganger"

10. Air travel
    a. Yes = “Reist med fly innenlands ELLER utenlands”
11. “Sociability”
    a. Low/medium = “Vært i folkemengde på 10-50 personer 3 eller færre ganger”
    b. High = “Vært i folkemengde på 10-50 personer 4 eller flere ganger”
12. Number of kids below 18 in the household
    a. Three categories: 0, 1-2, and 3 or more
13. Housing conditions
    a. Two categories: leilighet/tomannsbolig, rekkehus/enebolig
14. Living in urban areas
    a. Yes = “By/Tettsted = Ja”
15. Workplace of partner
    a. Health = “Helse med direkte kontakt OR Helse uten direkte kontakt”
    b. Store = “Butikk”
    c. Teacher = “Lærer/barnehage”
    d. Other = All other
16. Contact with people through work
    a. Yes = “Direkte kontakt med mennesker”
17. Pets
    a. Two categories: dog/cat, no pet/other
18. Previous light infections (mostly upper respiratory infections) past 6 months
    a. Yes = 1 or more infections
19. Handwash/use of protective equipment outside work
    a. Two categories: 0-6 times pr day, 7+ times pr day
20. Use of protetctive equipment outside work
    a. Yes = “Munnbind hver gang/noen ganger” ELLER “Hansker hver gang/noen ganger”
21. Time period for testing
    a. Three categories: Before March 13th/13th - 25th March/ after March 25th
22. Age
23. Sex

Some variables are “measured” both prior to the Norwegian lockdown on the 13th of March 2020, and after the lockdown. These are shopping habits, travel habits, and sociability. The rest of the variables are measured at the time the questionnaire was answered. Due to the biology of the virus, we expect that the variables regarding behavior after March 13 will not influence the results recorded until 1-3 weeks later. Therefore these will not be included in the analyses.

#### Sensitivity analyses

To assess robustness of results some sensitivity analyses will be done:

1. Excluding all those reported travelling to Italy and Austria

2. Looking at the subgroup of healthworkers only, since they have been exposed for a different testing regime.

3. Other sensitivity analyses might be performed depending of what we learn from data.

#### Rationale for defining different study populations

P1 cases and control group 1 come from a clearly defined base population – three main laboratories. Assuming 100% sensitivity and specificity of the test, they are all presumably truly cases and controls. The volunteer control group comes from a larger and undefined population in Norway.

P2 cases and controls come from an undefined larger population of volunteers in Norway. These are self-reported cases and controls. We have data on how good COVID-19 self reporting is from the P1 population where a very high proportion of participants with one test self-reported their test result in accordance with data from the laboratory (kappa >0.99), thus we presume self reporting is very good.

A challenge is that the P1 (and likely P2) cases and controls all had to satisfy *the criteria* for being COVID-19 tested. These changed over time (see above), and have been different for health care personnel than the population at large, but in general means that many of the P1 cases and controls will have had one or more symptoms associated with COVID-19 disease at the time of being tested. This means that *symptomatic P1 (and likely P2) controls* may have had another upper respiratory infectious disease such as the common cold or influenza-like disease. Behavior prior to testing can therefore have been similar between cases and controls (contact with sick person etc).
